# Assessing the burden of COVID-19 amongst healthcare workers in Mexico City: A data-driven call to action

**DOI:** 10.1101/2020.07.02.20145169

**Authors:** Neftali Eduardo Antonio-Villa, Omar Yaxmehen Bello-Chavolla, Arsenio Vargas-Vázquez, Carlos A. Fermín-Martínez, Alejandro Márquez-Salinas, Julio Pisanty-Alatorre, Jessica Paola Bahena-López

## Abstract

**BACKGROUND:** Health-care workers (HCWs) could be at increased occupational risk for SARS-CoV-2 infection due to increased exposure. Information regarding the burden of COVID-19 epidemic in HCWs living in Mexico is scarce. Here, we aimed to explore the epidemiology, symptoms, and risk factors associated with adverse outcomes in HCWs in Mexico City.

**METHODS:** We explored data collected by the National Epidemiological Surveillance System in Mexico City, in HCWs who underwent real-time RT-PCR test. We explored COVID-19 outcomes in HCWs and the performance of symptoms to detect SARS-CoV-2 infection.

**RESULTS:** As of September 20^th^, 2020, 57,758 HCWs were tested for SARS-CoV-2 and 17,531 were confirmed (30.35%); 6,610 were nurses (37.70%), 4,910 physicians (28.0%), 267 dentists (1.52%) and 5,744 laboratory personnel and other HCWs (32.76%). Overall, 2,378 HCWs required hospitalization (4.12%), 2,648 developed severe COVID-19 (4.58%), and 336 required mechanical-ventilatory support (0.58%). Lethality was recorded in 472 (0.82%) cases. We identified 635 asymptomatic SARS-CoV-2 infections (3.62%). Compared with general population, HCWs had higher incidence, testing, asymptomatic and mortality rates. No individual symptom offers adequate performance to detect SARS-CoV2. Older HCWs with chronic non-communicable diseases and severe respiratory symptoms were associated with higher risk for adverse outcome; physicians were at higher risk compared with nurses and other HCWs.

**CONCLUSIONS:** We report a high prevalence of SARS-CoV-2 infection in HCWs in Mexico City. Symptoms as a screening method is not efficient to discern those HCWs with a positive PCR-RT test. Particular attention should focus on HCWs with risk factors to prevent adverse outcomes.

## INTRODUCTION

The pandemic caused by the SARS-CoV-2 has created new challenges in health-care systems worldwide (1). Recently, the pandemic has led to significant increases in the number of cases in the Americas, where it has caused considerable pressure on health-care facilities and led to a substantial number of deaths (2,3). Health-care workers (HCWs) have a fundamental role in caring and managing patients with COVID-19, as they are the primary workers involved in the daily management of pandemic at an individual level. Notably, this population is at a significant occupational risk of infection attributable to an increased frequency of exposure to infected individuals, either symptomatic or asymptomatic, ultimately leading to an increased risk of associated COVID-19 complications. Although it has been emphasized that HCWs with respiratory symptoms should be isolated as soon as possible, there is no consensus on which other symptoms are suggestive of SARS-CoV-2 infection (4). Moreover, reports show that HCWs tend to present without respiratory symptoms at the time of diagnosis (5). Ultimately, this has raised questions on whether a testing strategy guided by the presence of symptoms and whether a mandatory policy of use of Personal Protective Equipment (PPE) outside the areas of care of infected patients are the best approaches in HCWs.

Additionally, the COVID-19 epidemic in Mexico coexists with a high prevalence of chronic health conditions, and HCW are no exception. Since such conditions have been amply shown to increase risk for severe outcomes in COVID-19, the pandemic creates a very complex situation (6). Furthermore, Mexico has a highly fragmented and segmented healthcare system, hampering its ability to provide a unified framework to protect HCWs (7). Besides, social inequities could have increase the disparities in risk among HCWs within marginalized communities (8). Given these fundamental differences, HCWs living in Mexico could be at a substantial risk for SARS-CoV-2 infection and adverse COVID-19 outcomes. There is a need to understand these trends and outcomes related to COVID-19 in HCWs to generate data-driven evidence which could inform public policy and promote development of recommendations to improve work environments and testing policies amongst HCWs by reducing viral transmission between peers and, ultimately decrease adverse outcomes. Here, we sought to investigate the epidemiology of SARS-CoV-2 infection within HCWs and its related outcomes in Mexico City.

## METHODS

### Data sources

We analyzed data collected within the National Epidemiological Surveillance System (NESS) database in Mexico City, which is an open-source dataset comprising daily updated suspected COVID-19 cases that have been tested using real-time RT-PCR to confirm SARS-CoV-2 according to the Berlin Protocol (9), and were certified by the National Institute for Epidemiological Diagnosis and Reference (10,11). The database contains information on all persons tested for SARS-CoV-2 infection at public facilities in Mexico, as well as in all private healthcare facilities that comply with the legal mandate to report COVID-19 cases to health authorities (12). Further testing strategy implemented in HCWs in Mexico City is presented in **Supplementary Material**. Data to estimate HCWs population rates were extracted from 2019 Health Information System obtained by the Mexican Ministry of Health (SSA) (13).

### Definitions of COVID-19 cases, predictors, and outcomes

Health-care related professions included subjects whose occupations were reported as physicians, nurses, dentists, laboratory personnel and other involved HCWs. Demographic and health data are collected and uploaded to the epidemiologic surveillance database by personnel from each corresponding health-care facility. Available variables include age, sex, nationality, state and municipality where the case was detected, immigration status as well as identification of individuals who self-identify as indigenous. Complete clinical assessment methodology is presented in **Supplementary Material**. Date of symptom onset, hospital admission, and death are available for all cases as are outpatient or hospitalized status, information regarding the diagnosis of clinical pneumonia, ICU admission, and whether the patient required mechanical ventilation support (MVS). Severe outcome was defined as a composite definition comprising death, requirement for MVS or ICU admission (14).

### Statistical analysis

#### Population-based statistics

Descriptive <u>analysis</u> methodology is presented in **Supplementary Material**. We estimated the incidence, testing, asymptomatic and mortality rates by standardizing each event among the reported 2019 HCW population working in Mexico City or 2020 population living in Mexico City using data from CONAPO (15), and normalized each estimate to reflect a rate per 100,000 inhabitants.

#### Conditions and symptoms related to SARS-CoV-2 positivity

We aimed to investigate comorbidities and symptoms associated with SARS-CoV-2 positivity using a mixed effects logistic regression model, including facility of treatment as a random effect to account for the variability in case distribution and treatment across healthcare facilities. We excluded HCWs who were suspected cases at the time of inclusion without a definitive result for SARS-CoV-2. Two separate models were designed to explore separately comorbidities and symptoms associated with SARS-CoV-2 positivity. We further explored the diagnostic test capacity, area under the curve, sensitivity, specificity, positive and negative predictive values (VPP, VPN, respectively) of each symptom to predict SARS-CoV-2 positivity.

#### COVID-19 mortality risk and clinical outcomes

We fitted Cox Proportional risk regression models to explore risk factors associated to COVID-19 related 30-day lethality, hospitalization, or severe outcome estimating time from symptom onset up to death, clinically reported outcome, or censoring (ongoing studied subjects), whichever occurred first. Factors associated with using mechanical ventilation support (MVS) were evaluated using a mixed effects logistic regression model. We also performed a Kaplan-Meier analysis to assess differences in COVID-19 outcomes comparing physicians and other HCWs using the Breslow-Cox test. A p-value <0·05 was considered as statistical significance threshold. All analyses were performed using R software version 3.6.2.

## RESULTS

### COVID-19 in health-care workers

As of the writing of this report (September 20^th^, 2020) the NESS had assessed for SARS-CoV-2 infection 403,185 people in Mexico City (estimated population: 8,737,172), out of whom 57,758 (14.32%) were HCWs (estimated HCWs population: 161,580). From the evaluated HCWs, 36.1% (n=20,848) were nurses, 30.8% (n=17,778) physicians and 33.1% (n=19,132) other-HCWs (5.51% dentists, 6.21% laboratory personnel and 88.28% supportive HCWs). Amongst them, 30.35% (n=17,531) had confirmed SARS-CoV-2 infection. Positivity rates by HCWs category were 31.2% (n=6,610) in nurses, 27.6% (n=4,910) physicians, 25.3% (n=267) dentists, 36.4% (n=433) laboratory personnel and 31.5% (n=5,311) for the supportive HCWs. In all studied HCWs, we recorded 2,648 (4.58%) HCWs who required hospitalization, 2,735 with a severe outcome (4.73%), and 336 who required MVS (0.58%). Lethality attributable was recorded in 472 (0.82%) HCWs. Overall, 635 (3.62%) HCWs referred no associated symptoms but tested positive for SARS-CoV-2 (**Table 1**).

**Table 1:** Characteristics amongst health-care workers assessed for SARS-CoV-2 test living in Mexico City. *Abbreviations*: COPD= Chronic obstructive pulmonary disease; HIV/AIDS= Human immunodeficiency virus and/or acquired immunodeficiency syndrome; CKD= Chronic kidney disease. *Other HCWs include: 1,056 (5.51%) dentists, 1,189 (6.21 %) laboratorians and 16,887 (88.28%) other related HCWs.

| Parameter | Physicians<br>(n=17,778) | Nurses<br>(n=20,848) | Other hcw*<br>(n=19,132) | P value |
| --- | --- | --- | --- | --- |
| Female sex (%) | 9,157 (5.15%) | 16,817 (80.6%) | 10,895 (56.94) | 0.001 |
| Positive cases (%) | 4,910 (27.61%) | 6610 (31.71%) | 6011 (31.42%) | 0.001 |
| Asymptomatic Cases (%) | 167 (3.40%) | 231 (3.49%) | 237 (3.94%) | 0.251 |
| Age (years) | 39.7 (11.9) | 37.97 (10.28) | 40.44 (10.93) | 0.001 |
| Mortality (%) | 211 (1.18%) | 64 (3.09%) | 197 (10.29%) | 0.001 |
| Hospitalization (%) | 1,059 (5.95%) | 739 (3.54%) | 850 (4.44%) | 0.001 |
| Severe Outcome (%) | 1,095 (6.16%) | 750 (3.59%) | 890 (4.65%) | 0.001 |
| Clinical pneumonia (%) | 1,233 (6.93%) | 1158 (5.55) | 1202 (6.28) | 0.001 |
| Mechanical-Ventilation (%) | 162 (0.91%) | 51 (0.24%) | 123 (0.64) | 0.001 |
| ≥7 days since beginning of symptoms (%) | 2,463 (13.85%) | 3417 (16.4%) | 3160 (16.51) | 0.001 |
| No-Comorbidities (%) | 11,696 (65.99%) | 13,295 (64%) | 11621 (60.97) | 0.001 |
| Diabetes (%) | 948 (5.34%) | 1,201 (5.77%) | 1162 (6.08) | 0.008 |
| COPD (%) | 105 (0.59%) | 69 (0.33%) | 103 (0.539) | 0.0004 |
| Asma (%) | 1075 (6.05%) | 675 (3.24%) | 574 (3.00) | 0.001 |
| HIV/AIDS (%) | 65 (0.37%) | 55 (2.64%) | 73 (0.382) | 0.086 |
| Obesity (%) | 2,164 (12.18%) | 3505 (16.85%) | 3245 (16.98) | 0.001 |
| Smoking status (%) | 1,270 (7.15%) | 1969 (9.46%) | 2495 (13.06) | <0.001 |
| CKD (%) | 106 (0.59%) | 79 (3.79) | 80 (4.18) | 0.0042 |
| Pregnant women (%) | 43 (0.24%) | 59 (2.83) | 56 (2.93) | <0.001 |
| Indigenous (%) | 55 (0.32%) | 73 (3.60) | 74 (3.96) | 0.4818 |

### Population-based analysis in health-care workers

Overall, we found that the positivity (30.3% vs 35.5%) and lethality rates (1.87% vs 9.20%) as well as daily confirmed asymptomatic cases (median IQR: 235 [181-307] vs 1,177 [324-3,047] cases) were lower in HCWs as compared with the general population. This can be partially explained by the fact that testing rates per 100,000 inhabitants were higher in HCWs compared to the general population (34,349 vs 3,825 per 100,000), which is also reflected in a higher incidence of confirmed (10,412 vs 1,359 per 100,000) and asymptomatic cases (3,728 vs 564 per 100,000). Nevertheless, in our population-based analysis we observed that the mortality rate (195 vs 125 per 100,000) was higher compared with general population living in Mexico City (**Figure 1**).

**Figure 1:**
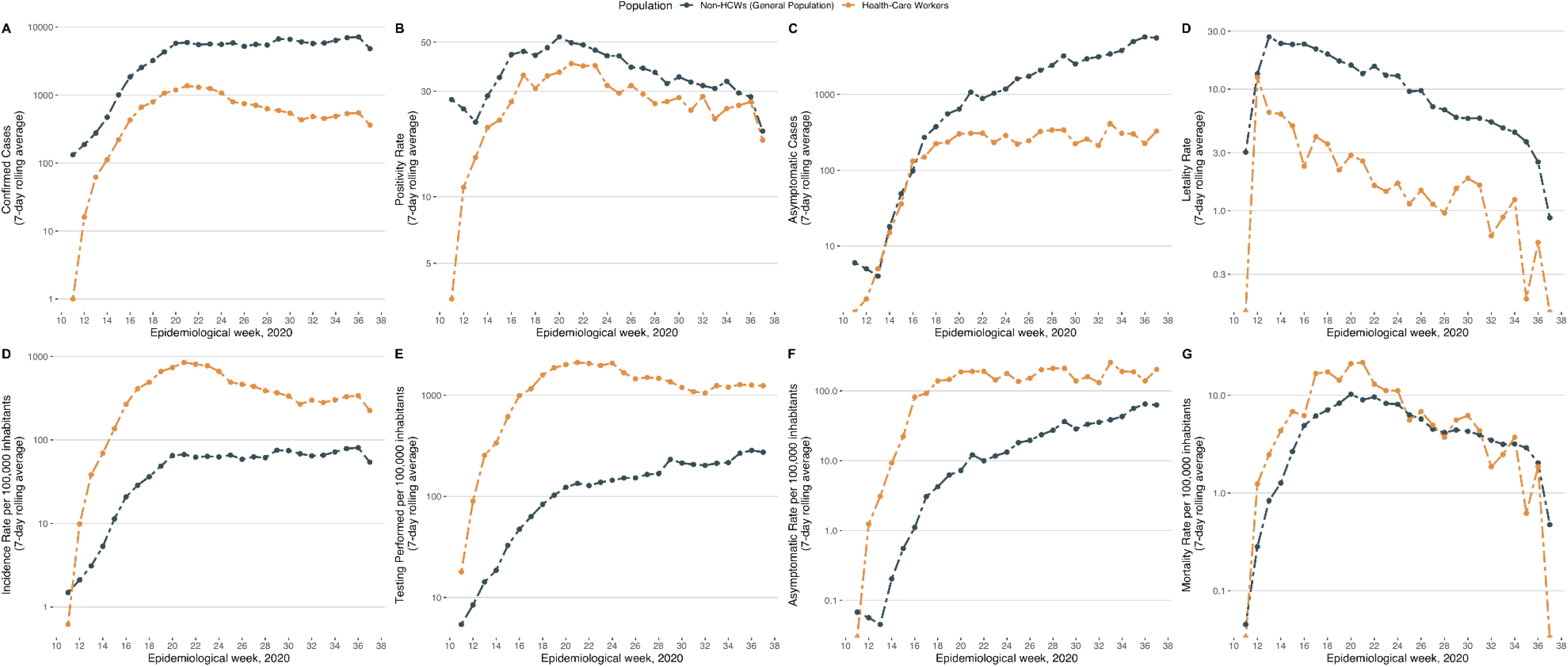
Confirmed SARS-CoV2 cases (A), positivity rate (B), lethality rate (C) alongside the incindence (D), testing (E) and mortality (F) rates per 100,000 inhabitants in health-care workers and general population with SARS-CoV-2 in Mexico City.

### Associated symptoms and conditions in health-care workers with SARS-CoV-2 positivity

Examining predictive performance of symptomatology to predict SARS-CoV-2 positivity, we observed that the presence of any symptom had an adequate sensitivity and negative predictive value, despite overall lower AUC. However, no individual symptom offered an adequate diagnostic accuracy or performance. Exploring the effectiveness to determine if known previous contact with suspected viral cases along with the absence of any symptom improved performance, we found that this joint consideration increased both sensitivity and negative predictive value, compared to only using the absence of any symptoms (**Supplementary Table 1**). Additionally, we found that male HCWs, with more than seven days from the onset of symptoms, with obesity, diabetes, or those pregnant women, had increased likelihood of SARS-CoV2 positivity; while those HCWs who reported active smoking, contact with a suspected viral case, with recent trips abroad and women in the puerperium period, had a lower probability for positivity (**Supplementary Figure 1**).

### Predictors for COVID-19 related outcomes in health-care workers

In confirmed SARS-CoV-2 cases, we found that symptoms at clinical assessment which increased risk for hospitalization were dyspnea, fever, and polypnea; while HCWs with diarrhea, odynophagia, and conjunctivitis had a decreased risk for this outcome. Furthermore, HCWs >65 years and those living with HIV/AIDS, diabetes, obesity, and arterial hypertension had increased risk of being hospitalized. Risk factors associated to severe outcomes in HCWs were age >65 years, living with diabetes, obesity and presence of dyspnea, fever, or polypnea at the time of initial clinical assessment. Moreover, those HCWs with age >65 years living with diabetes, early onset diabetes (<40 years), obesity, and seven days from the onset of symptoms or dyspnea at evaluation were conditions associated with MVS. Finally, predictors for lethality in HCWs were age >65 years, self-reported indigenous ethnicity, diabetes, early onset diabetes, obesity and presence of clinical pneumonia at evaluation (**Figure 2**).

**Figure 2:**
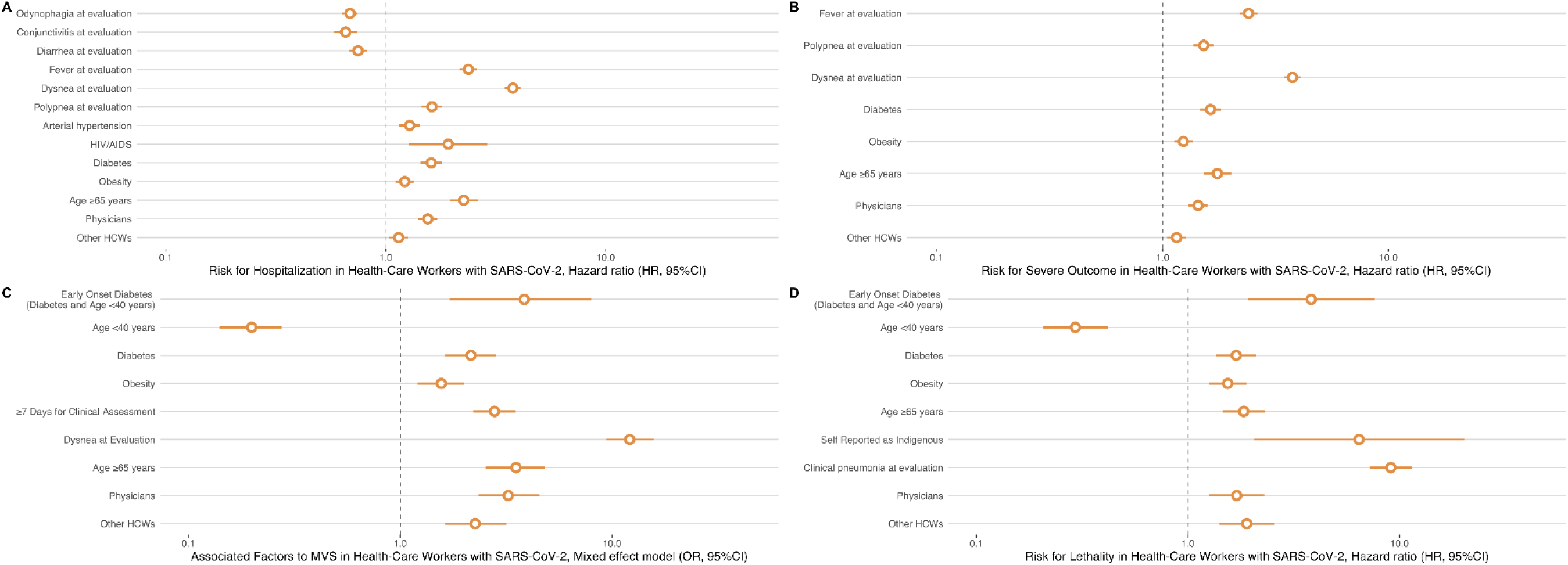
Risk factors associated to hospitalization (A), severe outcome (B), MVS (C) and lethality (D) associated to COVID-19 in health-care workers with SARS-CoV-2 in Mexico City. Abbreviations: MVS= Mechanical ventilation support.

### COVID-19 related outcomes of amongst groups of health-care workers

Overall, compared with general population with positive SARS-CoV-2 test living in Mexico City, HCWs had a decreased risk for hospitalization (HR 0.41, 95%CI 0.39-0.44), severe outcome (HR 0.46, 95%CI 0.44-0.49) and lethality (HR 0.33, 95%CI 0.30-0.37) (**Figure 3**). As a secondary analysis, we sought to explore risk of COVID-19 related outcomes in subgroups of HCWs. Although the group of physicians had a decreased likelihood to have a positive SARS-CoV-2 test (OR 0.81, 95%CI 0.77-0.85) compared with the group of nurses and other HCWs, this group had an increased risk for hospitalization (HR 1.57, 95%CI 1.36-1.81) and severe outcome (HR 1.34, 95%CI 1.17-1.54) after adjusting for the previously observed associated variables for each outcome compared with the group of nurses and other HCWs. We also observed that physicians had an increased lethality risk (HR 1.61, 95%CI 1.12-2.32) after adjusted for covariates (**Figure 3**). Finally, a particular group of interest were HCWs who belonged to an indigenous group, which had up to 14-fold increased risk for COVID-19 lethality (HR 95%CI 4.22-43.06) after adjusting for covariates, although no differences were found in the risk for hospitalization and severe outcome.

**Figure 3:**
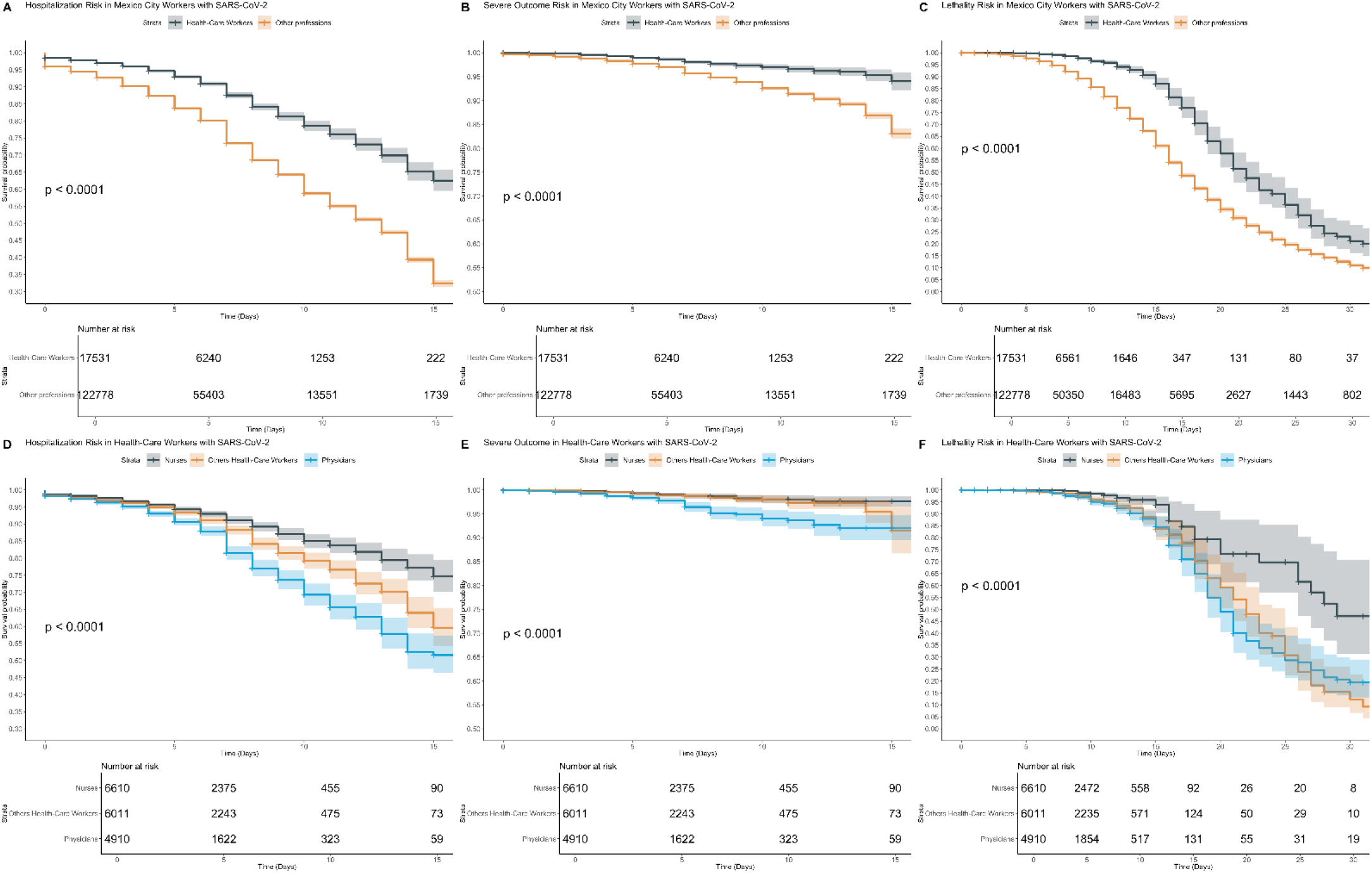
Kaplan-Meier survival plot to identify the risk of hospitalization (A-D), severe outcome (B-E) and lethality (C-F) in HCWs compared with other non-HCWs (general population) and between physicians and nurses and other HCW.

## DISCUSSION

In this work, we report the prevalence of SARS-CoV-2 infection, estimated epidemiological rates, and COVID-19 clinical outcomes using an open-source city-wide based surveillance reports of HCWs living in Mexico City. Amongst health-care workers, the group of physicians tend to have an increased risk of severe COVID-19 outcomes, which is a remarkable occupational risk. Moreover, certain factors, such as associated comorbidities and symptoms at the time of evaluation are associated with an increased risk of adverse outcomes. Particularly concerning is the fact that population-based mortality rate is higher among HCWs than the general population. It is unclear whether this reflects a true higher lethality, higher rates of infection, or a combination of both. These findings should be considered by authorities concerning the occupational hazards in HCWs, particularly in physicians to inform public policy related to ameliorate the burden of the COVID-19 pandemic on HCWs given their vital role in the daily pandemic management at an individual level.

HCWs have increased occupational hazard to acquire SARS-CoV-2 infection compared with general population, attributable to direct contact during care of hospitalized patients. Prevalence and lethality rates of SARS-CoV-2 infections reported in our HCWs population are higher compared with previous reports in China, Europe and the United States (4,16–22). Similarly, in Mexico the number of positive cases reported in HCWs has consistently increased since the first confirmed cases were reported in late February. To reduce transmissibility and improve outcomes, the current policy in Mexico requires that HCWs who develop respiratory symptoms should be isolated to mitigate the spread between peers. However, as our results confirm and based on previous reports, there is no sustainable evidence which allows using any individual symptom to identify SARS-CoV-2 positive cases in a clinical context (4). Nevertheless, a case definition of possible SARS-CoV-2 which includes the presence of any one symptom has acceptable sensitivity and a high negative predictive value, which is further improved by adding contact with symptomatic persons to the definition. Whilst the use of this definition may not eliminate the necessity to extend testing within all reasonable means, it does indicate that adequate contact tracing and case selection would allow to allocate testing in limited resource settings. Furthermore, the testing strategy adopted by the epidemiological surveillance system in Mexico, which is based on the appearance of symptoms to rule out diagnosis, may not be adequate to capture HCWs with asymptomatic infection or with non-respiratory symptoms. An alternative approach would be to include massive testing using low-cost detection tests for SARS-CoV-2 infectivity, which have recently been shown to have adequate sensitivity and could reasonably be implemented periodically for epidemiological surveillance amongst HCWs once such tests become widely available (23–25).

It is important to note that our results do not distinguish between HCWs directly caring for COVID-19 patients and those carrying out other tasks. It is therefore impossible to ascertain whether the higher COVID-19 risks relate to direct care or rather to high rates of public contact. Although previous reports have shown the effectiveness of personal protective equipment, there is still insufficient evidence if this strategy could mitigate the infection amongst all HCWs at long-term, which sets an area of opportunity for further studies (26,27). Based on the precautionary principle, we advocate a rational and equal access distribution of PPE in HCWs in Mexico City.

Our results also show that comorbidities in HCWs, particularly those related to chronic non-communicable diseases (e.g., diabetes, early onset diabetes, obesity and arterial hypertension), and the presentation of severe respiratory symptoms at the time of clinical assessment, increases the risk of adverse COVID-19 outcomes. Certain groups, such as pregnant women, older workers, and those who self-identify with indigenous ethnicity, are at higher risk for severe COVID-19 outcomes. Previous reports by our group had shown the relationship between the presence of cardiometabolic diseases and risk of complications associated with SARS-CoV-2 infection in Mexico (28–31). Although not completely understood, this relationship could be explained by immunological over responsiveness observed in confirmed cases with diabetes and obesity (28,31,32), particularly given recent evidence relating changes associated with an enhanced immune response to SARS-CoV-2 with risk of respiratory and multi-organ failure (31,33). Conversely, in patients with prior immunosuppression, an increased risk for associated co-infections has been reported, which could explain risk for adverse outcomes in HCWs with HIV/AIDS or pregnancy (34,35). Finally, we found that physicians are a group at risk for developing adverse events compared to other HCWs. This is consistent with a previous report, as it has been shown that physicians tend to spend more time in areas where patients with SARS-CoV-2 are assisted (36). Furthermore, prolonged shift times, work overload, psychological distress, exposure to probable cases amongst peers could lead physicians to be considered a group with significant occupational risk for developing COVID-19 related outcomes (37–39). Interestingly, we also found that groups of physicians who self-reported as belonging to an indigenous community were at increased risk of death attributable to COVID-19. Although preliminary, these results may denote an inequality in access to timely care in this group of HCWs, given the significant social discrepancies reported in Mexico, which have been exacerbated during the epidemic (8,40–42). More studies should focus on the risk of adverse outcomes attributable to social conditions in medical personnel.

Our study had some strengths and limitations. First, we analyzed a large dataset which included information on confirmed positive and negative SARS-CoV-2 cases in Mexico City, providing a unique opportunity to investigate COVID-19 specific risk factors in HCWs. A potential limitation of this study is the use of data collected from a sentinel epidemiological surveillance system model, which is skewed towards investigating high-risk cases based on the presentation of respiratory symptoms or only those with specific risk factors, which on the one hand increases power to detect the effect of comorbidities and on the other hand might not be representative of milder COVID-19 cases. This is particularly true for asymptomatic cases amongst HCWs, which were heavily underrepresented in our study and its prevalence must be assessed with widespread systematic testing amongst HCWs to reduce in-hospital transmission amongst peers. Although the testing performed in our HCWs population is higher compared to the general population living in Mexico City, it still remains low compared to other HCWs studies performed in other countries (43). Another potential limitation of our study is that our populational-based analyses of HCWs is limited to those living in Mexico City, without the exact number of recruited HCWs during the epidemic, thus our epidemiological rates should be interpreted as approximations. Moreover, all analyses were performed to estimate the burden of HCWs in Mexico City, which may not capture the whole picture in the country or the large socio-economic inequalities which might lead to higher rates of infection amongst HCWs in disadvantaged areas.

In summary, we present the first report of a city-wide based surveillance system which assessed clinical symptomatology and related outcomes attributable to COVID-19 in HCWs living in Mexico City. We found that no individual symptom can accurately discern HCWs with SARS-CoV-2 infection; therefore there may be a considerable but underreported prevalence of positive asymptomatic infections amongst HCWs. Symptoms should not be considered as the main indicator of infection and therefore, we strongly suggest a novel reframing for the current testing strategy implemented for HCWs living in Mexico City. Additionally, comorbidities, presence of respiratory symptoms at clinical assessment, and susceptible groups of HCWs, could have increase the risk of severe outcomes. Our results could inform policies within the health-care systems on a systematic testing policy, the rational use of personal protective equipment, early isolation of probable cases regardless the symptoms, exclusion of risk groups in areas where patients with SARS-CoV-2 are routinely assisted and consideration of intrinsic inequalities between workers, which overall, could bring to a better quality of life for HCWs during the COVID-19 pandemic.

## Data Availability

All data sources and R code are available for reproducibility of results at https://github.com/oyaxbell/covid_hcws_mx.

## ACKNOWLEDGMENTS

NEAV, AVV, CAFM, AMS, JPBL are enrolled at the PECEM program of the Faculty of Medicine at UNAM. NEAV, JPBL and AVV are supported by CONACyT. The authors would like to acknowledge the invaluable work of all of Mexico’s healthcare community in managing the COVID-19 epidemic. Its participation in the COVID-19 surveillance program has made this work a reality, we are thankful for your effort.

## AUTHOR CONTRIBUTIONS

Research idea and study design: NEAV, OYBC; data acquisition: NEAV, OYBC; data analysis/interpretation: NEAV, OYBC, AVV, CAFM, AMS, JPA,JPBL; statistical analysis: NEAV, OYBC; manuscript drafting: OYBC, NEAV, AVV, CAFM, JPA, AMS, JPBL; supervision or mentorship: OYBC. Each author contributed important intellectual content during manuscript drafting or revision and accepts accountability for the overall work by ensuring that questions about the accuracy or integrity of any portion of the work are appropriately investigated and resolved.

